# Closed-Loop Quality Assurance for Production Clinical AI Documentation

**DOI:** 10.64898/2026.05.27.26353977

**Authors:** Andrew Napier, Justin Wiley, Mark Heslin

**Affiliations:** Stanford School of Medicine. Stanford University, Stanford, CA, USA; Sayvant Health. San Francisco, CA, USA

**Keywords:** clinical AI, autonomous prompt optimization, closed-loop QA, deterministic post-processing, multi-agent pipelines, recurring local validation, LLM-as-judge

## Abstract

A closed-loop quality system deployed across thirteen US hospital sites resolved physician complaints with zero regressions on 42 tracked cases across 1,089 optimization iterations, while a deterministic assembly-agent replacement cut H+P trace latency from 19.6 s to 10.8 s (−8.8 s, 95% CI [−10.5, −7.1] s; *n* = 100 pre, *n* = 100 post). The system autonomously diagnoses, repairs, and verifies quality in production clinical AI documentation across academic medical centers, large urban community hospitals, mid-size regional hospitals, and small community hospitals. The 1,089-iteration operational metrics from a 36-day window (March–April 2026) predate the three-axis Pareto commit rule described in §3; Pareto-era operation continued through May 2026, and we report the architectural follow-through but defer comparable longitudinal keep-rate evidence to a future report.

We report four observations from operating this system over 36 days across 66 agents, plus the architectural follow-through that arrived six weeks later and provides operational evidence for the most contestable of the four. First, we hypothesize a distinction, supported by evidence from one pipeline, between *reasoning agents* (those generating new clinical content from context) and *assembly agents* (those arranging upstream outputs into a final format). In our pipeline, prompt optimization worked on the reasoning agents but did not work on the assembly agent: four consecutive optimization attempts on one assembly agent produced 18–28 point regressions. The effective intervention for assembly agents is deterministic post-processing. We initially deployed 166 such rules across 6 profiles; in a controlled ablation on 25 cases (*n* = 25, drawn from the multi-site case pool), these rules improved binary check pass rate by 11.7 percentage points and eliminated all 6 fabrication-class failures. **The architectural follow-through (§6): six weeks after the prediction was made, we replaced the assembly LLM call entirely.** The Chart_Builder agent on the hospitalist pipeline was migrated from a multi-kilobyte LLM prompt to a sub-500-character deterministic template plus sandboxed scripting that arranges the upstream agent outputs. Deterministic post-processing was not the endpoint of the architectural argument; it was a way station. The endpoint was deleting the LLM call.

Second, the same binary check instrument produces opposite outcomes depending on how it is used: as an optimization target, it yields notes scoring 90%+ that physicians reject (*ρ* = −0.077, *p* = 0.652, *n* = 36; not statistically significant); as a deployment gate asking “did this specific fabrication stop?”, it provides rater-invariant commit decisions.

Third, a calibration study expanding from one physician rater to three (*n* = 84 ratings across 36 blind pairs) found that the physician-preference signal is rater-fragile at this sample size: between two board-certified emergency physicians, Cohen’s *κ* = 0.028 with 95% CI [−0.30, 0.36] on *n* = 35 overlapping pairs. The CI’s lower bound reaches zero on all three rater pairs; the data are consistent with no agreement above chance and do not establish ordered calibration across raters. The structural finding is that physician preference cannot be treated as a single optimization target at this scale. We define quality operationally as: absence of fabrication (binary checks), structural completeness (deployment gates), and per-physician preference tracking (pairwise monitoring). Neither objective checks nor subjective preference alone is sufficient.

## 1 The Quality Maintenance Problem

Clinical documentation AI converts physician dictations into structured medical notes through pipelines of 6–19 specialized agents. A typical admission note flows through a routing agent, a summarizer that extracts structured data from the dictation, four narrative agents that generate the history, physical exam, and assessment/plan sections, and an assembly agent that merges everything into the final document. The output must simultaneously satisfy clinical accuracy (no fabricated findings), billing compliance (correct diagnostic specificity), and physician acceptance (the doctor would sign it). We report from a multi-site production deployment spanning thirteen US hospitals, a mix of academic medical centers and community hospitals of varying sizes, collectively generating notes daily across hospital medicine and emergency medicine.

Errors compound across agents. A medication misclassified by the extraction agent propagates through summarization, narrative generation, and final assembly. A fabricated pertinent negative (“denies nausea”) inserted by one agent gets treated as clinical fact by every downstream agent.

Manual review does not scale. Each hospitalist site generates hundreds of notes daily; aggregated across the thirteen sites in this deployment, the daily volume is in the thousands. Each note passes through multiple agents, each capable of introducing fabrications, omissions, or formatting errors. The review burden grows linearly with volume and agent count.

The operational question is not “how good is the AI?” but “who maintains quality after deployment?” Health systems need three capabilities: detection (find problems), diagnosis (identify which agent caused them), and repair (fix the root cause without breaking something else). Today, these require human prompt engineers reviewing individual cases. This gap between technical AI performance and operational outcomes has been characterized as requiring a separate “delivery science” for clinical AI [Shah, 2020]. Our system automates detection, diagnosis, and repair, because the quality of clinical AI documentation directly affects patient care.

### Deployment

The system reported here is deployed across thirteen US hospital sites, spanning the range of US inpatient care settings: academic medical centers, large urban community hospitals, mid-size regional hospitals, and small community hospitals. The deployment covers hospital medicine (HM) and emergency medicine (EM) note generation across these sites; aggregated daily note volume is in the low thousands. All sites use mainstream commercial EHRs and commercial ASR-vendor dictation pipelines whose output is consumed by the Sayvant Health multi-agent documentation system described below. Specific site identities are withheld for confidentiality (§11). The operational metrics reported here aggregate across the thirteen-site footprint over the 36-day window (March–April 2026), plus the F17 architectural follow-through deployed in May 2026 on the hospitalist admission profile (which serves all thirteen sites). The multi-site footprint provides natural variation across patient population, payer mix, physician staffing model, and case complexity; replication *outside* this footprint (international, federal/VA, pediatric-only institutions) is listed in Limitations (§10).

### Contributions of this paper

We make five contributions, summarized here for the reader’s roadmap:

1. **Closed-loop QA architecture** (§3, §4): a three-layer system, deterministic server-side repair, binary-check deployment gates, pairwise-preference monitoring, that detects, diagnoses, and repairs quality regressions in deployed clinical documentation AI without human prompt engineers in the loop. Operationalizes “recurring local validation” [Shah et al., 2023].
2. **Pareto-with-absolute-floors acceptance rule** (§3): a multi-axis commit rule with severity-class categorical vetoes, articulated as an alternative to scalar-reward acceptance for safety-critical autonomous optimization.
3. **Reasoning/assembly agent distinction** (§5.2, with construct-validity caveat in §5.2.1): a hypothesized distinction between multi-agent pipeline stages that respond to prompt optimization (reasoning) and those that do not (assembly), with three falsifiable predictions registered.
4. **F17 architectural follow-through** (§6; see Appendix D for the “F17” nomenclature): six weeks after the assembly-agent observation, the LLM call at the chart-assembly step was replaced with a deterministic renderer (compact template plus sandboxed scripting). The architectural endpoint of the assembly-agent thesis was deletion of the LLM call, not better wrapping of one.
5. **Cross-iteration rejection memory** (§3.5): a pattern that surfaces edit signatures rejected or reverted ≥ 3 times across iterations to the optimizer’s prompt, preventing the loop from re-proposing pre-losing edits.

## 2 A Worked Example

A physician flagged a note. The following is a synthetic reconstruction preserving the fabrication pattern while removing identifying details:

*“The dictation contained no mention of goals-of-care discussions or code status. The generated note included:* Patient and family have discussed goals of care. Patient expressed desire to avoid aggressive measures including intubation and resuscitation. Family member is designated healthcare proxy. Code status: DNR/DNI.*”*

The AI had fabricated an entire goals-of-care conversation. It invented a family member as healthcare proxy, generated a do-not-resuscitate order, and attributed end-of-life preferences that were never discussed, none of which appeared in the physician’s dictation. This category of hallucination, fabricating a patient’s end-of-life wishes, represents the highest-stakes failure mode in clinical documentation.

72 hours from complaint to verified production fix. No human prompt engineer involved. The deterministic repair rule catches any future fabricated code status at zero token cost. The prompt gate prevents the model from generating it. The regression gate ensures the fix is never broken by future optimization. This loop runs every night.

## 3 System Architecture

### 3.1 Layer 1: Deterministic Server-Side Repair

Per-agent scripts execute after each LLM call, operating on raw output plus the original dictation. Zero additional tokens. Negligible latency (*<*5 seconds total per note). At the time of the headline metrics (§8, March–April 2026) the system maintained 166 rules across 6 deployed profiles spanning six categories (Table 1). The registry is actively curated: weekly pruning removes redundant or dead rules and weekly additions target newly cataloged failure patterns. §6 reports the May 2026 architectural follow-through that converted one assembly agent from an LLM call with repair rules into a deterministic renderer with no LLM call at all.

**Table 1:** Deterministic repair rule categories at the April 2026 snapshot.

| Category | Example | Count |
| --- | --- | --- |
| Fabrication | “denies nausea” when nausea never mentioned | 85 |
| Omission | ED medications in dictation but missing from output | 21 |
| Format | Section breaks destroyed by agent handoff | 34 |
| Terminology | “heart attack” instead of “myocardial infarction” | 9 |
| Consistency | Female pronouns when dictation says male | 5 |
| ASR repair | “lays six” → “Lasix” | 12 |

Rules are written in a sandboxed Python subset that runs server-side on every note generation. Each rule has access to the agent’s output, the original dictation, and upstream agent outputs, enabling cross-reference checks that the generative model cannot perform.

In head-to-head evaluation on six test cases, deterministic repair eliminated all fabrication-class failures (6 → 0). On one representative case, adding 25 new repair rules in a single session moved physician-assessed note quality from 5/10 to 9.5/10, an observation from a single case that should not be generalized without broader evaluation.

### 3.2 Layer 2: Binary Check Deployment Gates

282 binary checks evaluate each candidate note. Category-specific evaluation follows evidence that aggregate LLM performance scores mask substantial variation across clinical task types [Chen et al., 2025]. Each check tests a single property: “Does the Plan section fabricate monitoring orders not in the dictation?” or “Is the encephalopathy diagnosis used only when altered mental status keywords appear in the source?”

The checks serve as deployment gates, not optimization targets. This distinction is critical (§5.1). The commit decision is a Pareto-frontier rule over three axes that we measure every iteration: (i) *binary target*, pass rate of the specific checks mapped to the active failure pattern; (ii) *narrative*, the mean of six narrative quality dimensions (story cohesion, clinical completeness, natural flow, absence of artifacts, physician readability, input fidelity); (iii) *pairwise preference*, the physician-calibrated A/B judge (§5.3). A candidate commits when it *dominates* the baseline (no axis worse, at least one strictly better) or is *non-dominated* (at least one strict win, at most one strict loss across measured axes).

Two absolute-floor reverts sit above the Pareto rule and fire regardless of the other axes:

1. **Gate-class violation.** Any check classified as gate-class (predominantly fabrication checks) that went from ≥ 1 passing case to zero passing cases.
2. **Major-severity regression.** Any check classified as major-severity (would change a clinical decision) that newly fails in the candidate relative to the baseline, or a net increase in major-severity failures. A formalized clinical-severity rubric (§3.3.1) extends this floor to pairwise-judged cases: any case where the baseline is preferred and the severity judge classifies the difference as major forces pairwiseOutcome = loss regardless of raw vote count.

Holdout validation is a separate gate that fires after commit: a post-commit run against cases the optimizer never sees during training, with divergence beyond 10% flagging overfitting. Hold-out data is accessed only through a dedicated module (§7.2) that refuses any path outside the holdout directory.

The Pareto rule replaces an earlier single-signal rule (“binary target must improve, overall failures cannot increase by more than 2”) that discarded candidates with flat binary target but strict narrative or pairwise wins. The three-axis formulation preserves those wins.

### Pareto-with-absolute-floors as a methodological contribution

Most autonomous prompt-optimization frameworks accept a candidate edit on a single scalar signal: a reward score (RLHF [Christiano et al., 2017, Ouyang et al., 2022], DPO, PPO), a metric improvement (DSPy [Khattab et al., 2023], OPRO [Yang et al., 2024]), or a self-critic verdict (Reflexion [Shinn et al., 2023], SELF-REFINE [Madaan et al., 2023]). Scalar acceptance is appropriate when the optimization target collapses cleanly to one dimension; it is misaligned with safety-critical applications where multiple distinct quality properties must be jointly maintained and where a single class of regression (here: fabrication; in other domains: factual error, harmful output, regulatory non-compliance) must be treated as an absolute constraint rather than a trade-off.

The architecture in this paper articulates a different acceptance rule: a Pareto-dominance check over *k* measured axes (*k* = 3 here: binary target pass rate, narrative quality, pairwise preference), gated above by a small set of absolute-floor constraints (gate-class fabrication, major-severity regression). Edits commit when they Pareto-dominate or are Pareto-non-dominated across the measured axes *and* clear every absolute floor. The absolute floors are not weighted into the Pareto comparison; they are categorical vetoes. We have not seen this exact decomposition, multi-axis Pareto acceptance with severity-class absolute-floor vetoes, articulated in the prompt-optimization literature, and we name it explicitly here because we believe it generalizes beyond clinical documentation to any safety-critical autonomous-optimization setting where the safety guarantee cannot be averaged against the quality signal. The cost of the rule, relative to scalar acceptance, is a lower keep rate (39% under the prior single-signal rule, expected to be lower under three-axis Pareto); the benefit is that no safety-class regression silently commits because it was offset by a quality gain on a different axis.

### 3.3 Layer 3: Pairwise Preference

A pairwise judge compares candidate output against baseline. One instruction: “Which note would you sign?” No rubric, no dimensions, no anchors. This minimal formulation achieved 61% agreement with one physician’s blind ratings. Three iterations of adding guidance all regressed: 47%, 53%, 52% (§5.4; full prompt texts in Appendix B). This represents single-point calibration rather than validated scalable oversight.

Pairwise contributes as the third Pareto axis and is computed every iteration on admission as of the 2026-04-18 AI council fix. Other note types remain on the prior sampling cadence pending stability verification on admission. Providing the clinician’s original dictation alongside both notes was the single largest improvement, without it, the judge cannot catch omissions or recharacterized findings.

#### 3.3.1 Clinical severity rubric

##### Operational status

The severity rubric described in this section is documented and versioned but is not yet enforced in the commit rule at the time of writing. The classification module and the severity-weighted accept routine are implemented; the feature flag wiring the severity judge into the live Pareto axis is off. The subsection describes the target architecture; the operational metrics reported in §8 pre-date the rubric and reflect the raw-vote formulation. A shadow-mode rollout is planned on admission only. The gating criterion to turn the feature flag on is two-part: (i) at least 30 shadow-mode classifications with ≥ 80% agreement against a physician-reviewed severity label on the same case, and (ii) zero major-class false negatives on a 12-case adversarial set drawn from previously-resolved patient-safety complaints. Both thresholds are pre-registered internally before the shadow run begins.

The raw pairwise vote (prefer-candidate vs. prefer-baseline counts) treats every case’s preference signal as equal weight. A clinical-severity rubric stratifies cases into three classes before the Pareto rule reads the axis: *major* (patient safety or liability risk: fabricated medications, demographic mismatch, hallucinated treatment), *minor* (clinical quality friction: missing pertinent negatives, weak clinical reasoning, incomplete differential), and *cosmetic* (formatting or stylistic preference). The full rubric is versioned internally; we describe its categories but do not release the per-class decision tree.

The rubric integrates with the pairwise axis in two ways. First, any case where the baseline is preferred and the severity judge classifies the difference as major forces pairwiseOutcome = loss regardless of raw vote count, generalizing the absolute-floor pattern already present for gate-class violations and TD regressions into a first-class severity axis. Second, minor-severity cases contribute normal weight (1.0) to the pairwise delta while cosmetic-severity cases contribute reduced weight (0.25), so a 10-case candidate-preferred result driven by header capitalization does not outweigh a single baseline-preferred case where the candidate has weaker clinical reasoning.

### 3.4 Runtime Gate Hierarchy

Each candidate edit traverses a staged gate hierarchy between the agent’s write and the commit decision. Gates are ordered cheapest-first so obvious breakage is caught before expensive runs:

1. **TypeScript syntax lint.** tsc –noEmit on the diff. Catches syntax errors in prompt files before any downstream cost.
2. **JSON validation.** JSON.parse on each edited profile JSON. Catches malformed Sayvant Guard profiles.
3. **Anti-pattern lint.** Regex scan of the diff against five known-bad patterns (e.g., cleanup_punctuation added to a JSON-outputting agent, or short-substring text_contains calls that match inside longer words). Each pattern corresponds to a prior production outage.
4. **Self-review.** A lightweight LLM critic reads the diff and the top-3 currently failing checks; rejects the edit if the change plausibly does not address those checks. Prevents scope-drift edits.
5. **Runtime smoke gate.** A 1-case end-to-end pipeline check runs against the modified prompts before the full measure batch. On failure, the working tree is reverted and the iteration is logged as a smoke-gate failure without burning the measure batch (∼10–30 min saved per bad edit).
6. **Measure.** The full grading batch (8–15 cases) generates notes through the production pipeline on staging and evaluates against the 282-check matrix. Produces the binary and narrative axes.
7. **Decide.** The Pareto rule determines commit or revert.

Figure 3 renders the gate sequence with the Pareto-rule decision node and the cross-iteration-rejection-memory feedback path that feeds future *Competitor* prompts (the LLM agent in step 3 of §4 that proposes edits; nomenclature used in the following subsections).

**Figure 1:**
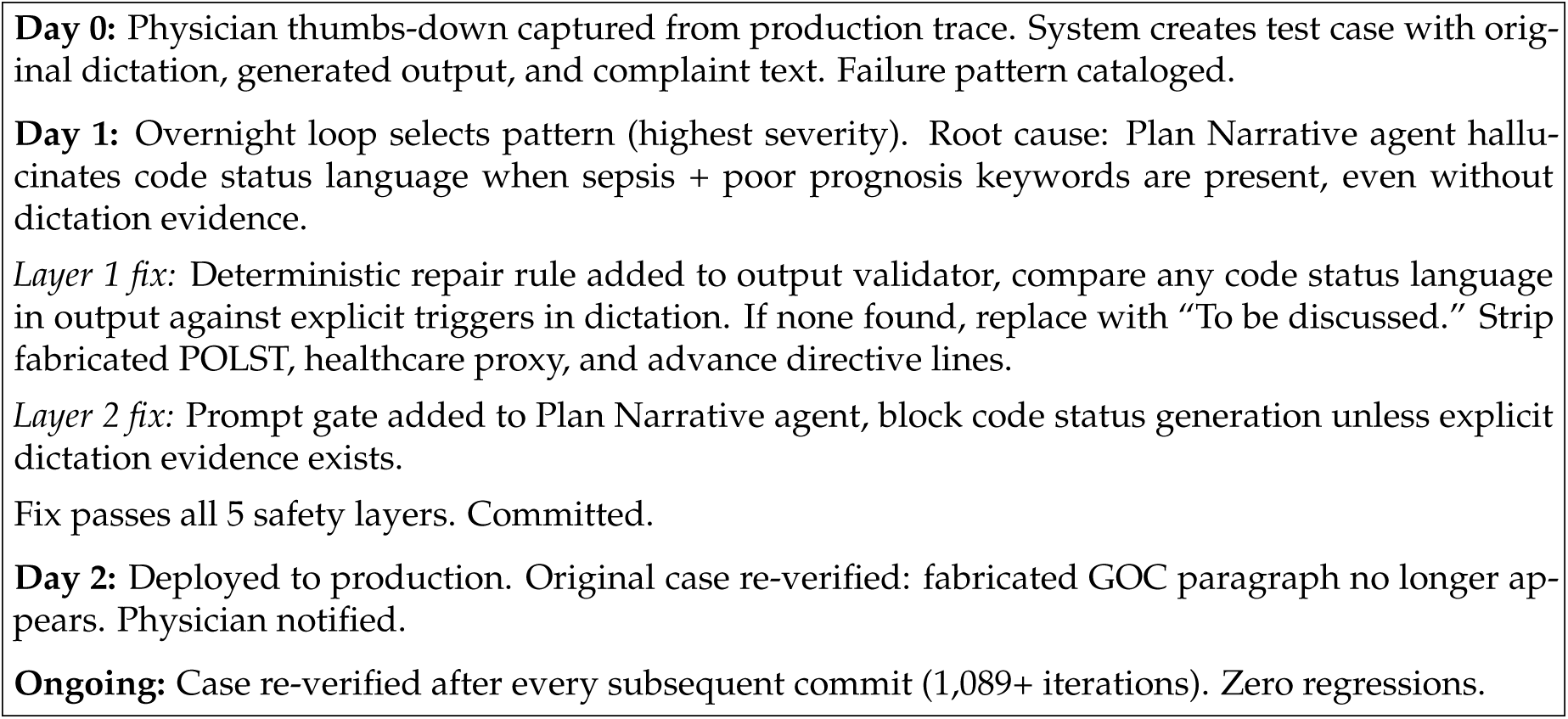
Complaint-to-fix timeline for TD-077: fabricated DNR/DNI order. The two-layer fix combines a deterministic repair rule (zero token cost, catches all future instances) with a prompt gate (prevents generation). Total time: 72 hours.

**Figure 2:**
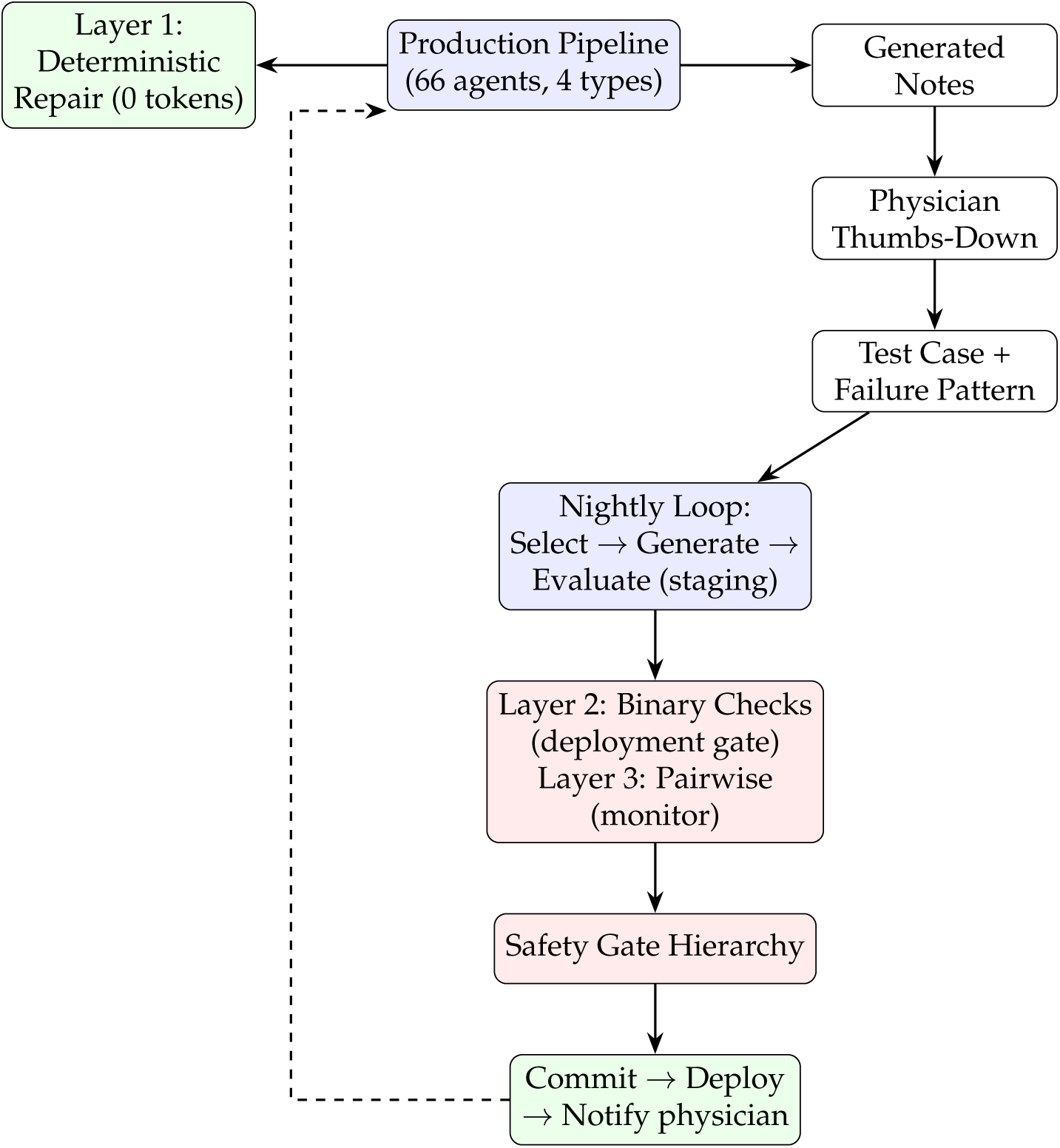
Closed-loop architecture. Physician complaints drive test case creation. The nightly optimization loop generates fixes, evaluates against binary checks and pairwise preference, and deploys through the safety gate hierarchy. The dashed arrow indicates production deployment completing the loop.

**Figure 3:**
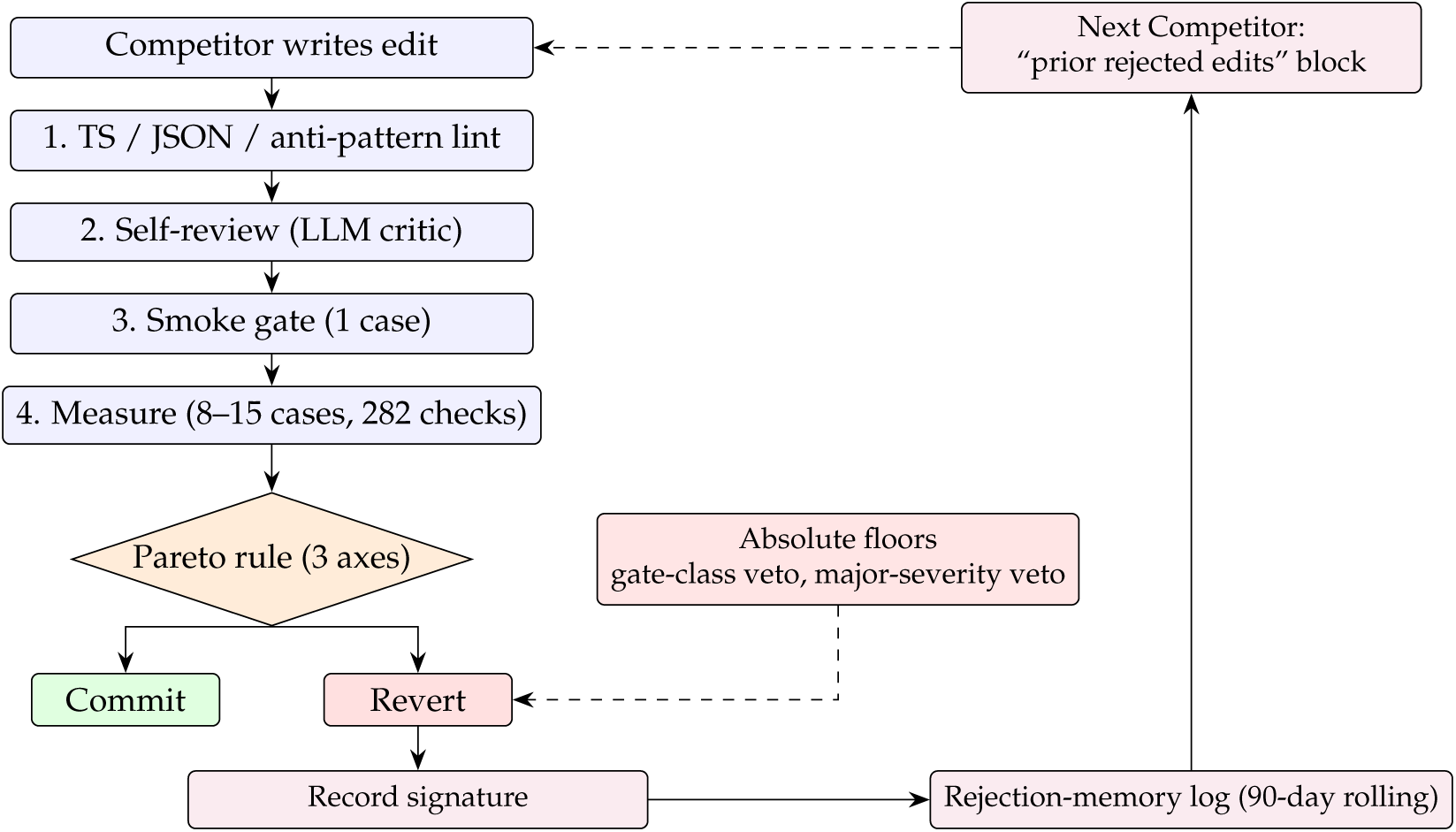
Gate-flow with Pareto decision and rejection-memory feedback. Solid arrows: forward path. Dashed arrows: categorical-veto (Absolute floors → Revert) and the cross-iteration feedback path. Absolute floors (gate-class or major-severity regression) force a revert regardless of Pareto axes. Every revert records an edit signature (cryptographic hash of changed files plus leading added lines) in a per-note-type log; signatures with ≥ 3 negative outcomes surface to the next Competitor as a “prior rejected edits” block, closing the loop on cross-iteration learning. Lint, self-review, and smoke-gate failures (§3.4) also write to the same signature log; their arrows are omitted from the diagram for legibility.

**Figure 4:**
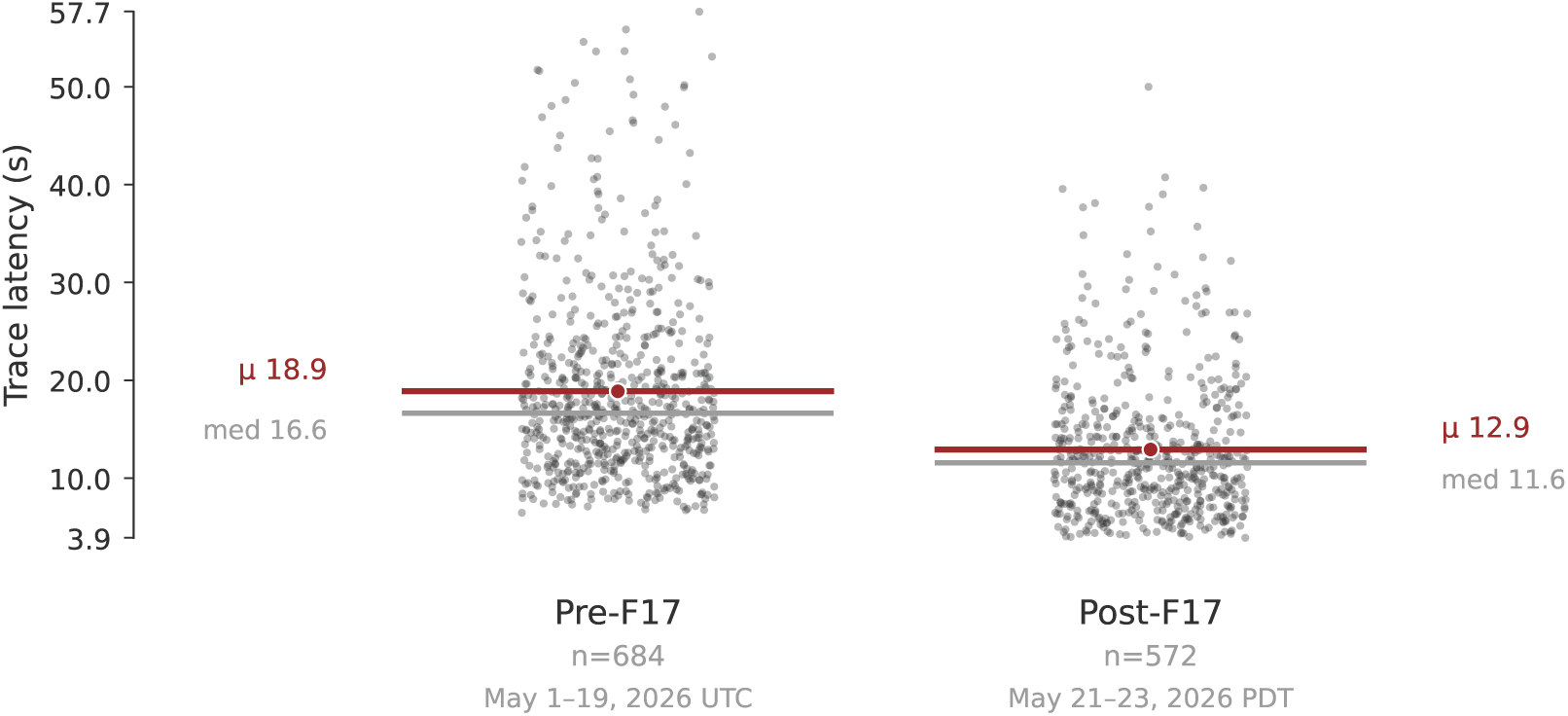
HM admission H+P trace latency, pre- vs post-F17, individual observations. Each dot is one Langfuse trace from the UAT controlled-batch pipeline (tag filter batch+hm2, name H+P), outlier-clipped at ≤ 60 s. Red bar marks the mean; gray bar marks the median. Pre-F17 window matches Table 4 (May 1–19, 2026 UTC; *n* = 684). Post-F17 is restricted to May 21–23, 2026 PDT (*n* = 572); the May 20 deploy day is excluded because its mean latency (18.3 s) is indistinguishable from pre-F17 days, indicating the change had not yet stabilized. Table 4 reports a smaller controlled *n* = 100/*n* = 100 sub-batch with slightly tighter selection criteria; the architectural pattern (lower mean, narrower distribution, compressed right tail) holds across both samples. Mean dropped 18.9→12.9 s in this larger sample (−32%); SD narrowed 9.4→7.1 s (−25%). Raw latencies released alongside the reproducibility bundle (§11).

### 3.5 Cross-Iteration Rejection Memory

The gate hierarchy described above rejects edits within a single iteration, but gave the optimizer no memory across iterations. A concrete failure mode surfaced on 2026-04-20: the Competitor proposed the same “FIX 8 HPI fabricated-negatives” edit on both the progress and discharge note types on the same night; both were rejected by self-review as CONFLICT with the existing denies-scan; and the denies-scan-removal edit pattern had accumulated 13+ prior rejections in per-type Curator history (the post-iteration analyst that distills commits and reverts into failure-pattern entries). None of that rejection history reached the Competitor at the time it was composing the next edit, the agent had no way to know it was replaying a pre-losing move.

We introduced a cross-iteration signature log adapted from a similar replay-protection pattern in published self-improvement-loop code. After every rejected or reverted edit, we record a short cryptographic-hash signature of the changed files plus the leading non-blank, non-comment added lines, together with the rejection outcome and a short reason, in a per-note-type log with a 90-day rolling window. Before the Competitor writes, we load that window’s signatures, group them, and surface any signature with ≥ 3 negative outcomes in the Competitor prompt as a “PRIOR REJECTED EDITS” block. The agent is instructed to either propose a materially different approach, commonly moving the fix to a different layer such as the safety net or down to deterministic post-processing, where the execution model tolerates verification rules that the narrative-generation prompts do not, or to explain in a mandatory diff-description line why this attempt differs from the prior ≥ 3. The implementation is a small library with recording hooks at all four in-iteration rejection sites and one post-measure revert site.

## 4 The Autonomous Loop

The system runs nightly. Each cycle:

1. **Harvest.** Pull new physician complaints and production traces. Convert to test cases with dictation, output, and complaint text.
2. **Select.** Rank failure patterns by impact. Physician complaints get priority. Each pattern maps to specific binary checks and a recommended fix type: deterministic rule, prompt edit, or both.
3. **Generate.** A Claude Opus agent receives the strategy document, failing check context with output/dictation snippets, iteration history, example diffs from prior successful commits, and, where applicable, the cross-iteration rejection memory block (§3.5) listing edit signatures rejected or reverted 3+ times. It edits prompt files or proposes new repair rules.
4. **Evaluate.** Generate notes for 8–15 test cases through the production pipeline on staging. Evaluate against the full 282-check matrix. Compare against baseline.
5. **Decide.** Commit if the candidate dominates or is Pareto-non-dominated across the three axes. Revert on gate-class violation or major-severity regression regardless of other axes. A 39% keep rate was measured under the prior single-signal rule (1,089 iterations, March–April 2026); Pareto-era keep rate is reported as ongoing.
6. **Validate.** Holdout cases the optimizer never sees during training. Extended validation after consecutive commits.
7. **Safety.** Multi-layer gates between optimizer and production (§7).

The loop provides the optimizer with raw per-case reasoning, not compressed scores. “Score dropped 2%” produces random fix attempts. The targeted form is a per-case failure record:

case_id: POS-42

failing_check: ap.no_fabricated_meds

agent: AP_CONDENSER

output_excerpt: “Continue enoxaparin 40mg SC daily for DVT prophylaxis.”

dictation_excerpt: [no anticoagulant mentioned]

upstream_state: AP_EXTRACTOR did not list enoxaparin; AP_CONDENSER inserted it.

last_5_iterations_on_this_check: [4 reverts, 1 no-op]

suggested_fix_type: deterministic_repair_rule (per playbook)

Full execution traces, not summaries. The per-iteration history block prevents the optimizer from re-proposing edit patterns it has already cycled through, complementing the cross-iteration rejection memory described in §3.5.

Over 36 days (March–April 2026): 1,089 iterations, 290 committed improvements, 316 autoresearch-related commits. The system maintains ∼40,000 lines of prompt code across 66 agents spanning four note types in two specialties. Nightly runs have continued through May 2026 with the architectural changes described in §6.

## 5 Key Observations

### Statistical methods (preamble to §5)

All inferential statistics reported in §5 and §8 are computed as follows. *Confidence intervals on proportions:* Wilson 95% CI [Wilson, 1927], computed analytically. *Confidence intervals on paired differences in proportions:* non-parametric bootstrap with 10,000 resamples on case-level pass rates, percentile method for the 95% CI. *Confidence interval on Spearman ρ:* Fisher *z*-transform with *n* – 3 degrees of freedom. *Confidence interval on Cohen’s κ:* the 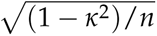 approximation, conservative at small *n*. *Software:* all CIs computed in Python 3.11 with seed 42 for bootstrap reproducibility; source script and per-case data are released in the public reproducibility bundle (§11). *Multiple comparisons:* this paper reports per-test 95% CIs and treats individual results as descriptive within the small-*N* regime; no family-wise adjustment is claimed (§10 elaborates).

### 5.1 Optimization Target vs. Deployment Gate

The same evaluation instrument produces opposite outcomes depending on how it is used.

#### As optimization target

39 structural checks. Notes scored above 90%. Spearman correlation against physician preference: *ρ* = −0.077 (95% CI [−0.396, 0.258]; *p* = 0.652, *n* = 36; method per §5). The CI spans both directions of correlation; we report this as directional evidence that high structural scores did not predict physician preference in our sample, not as a precise measurement. At this sample size we cannot rule out a modestly positive or modestly negative correlation, only that the data do not support a strong positive one.

#### As deployment gate

282 checks including fabrication and clinical quality. Question changed from “increase overall score” to “did this specific fabrication stop?” The first conflates structure with substance. The second is verifiable and rater-invariant.

#### Registered replication

At *n* = 120 calibration pairs against the majority vote of ≥ 3 raters, we predict (i) |*ρ*| between high structural scores and physician preference remains below 0.2, and (ii) rater-invariance on the gate-class subset holds with variance across raters below 0.05. Timeline: Q3 2026 calibration expansion. If |*ρ*| *>* 0.4 at *n* = 120, the target-vs-gate distinction is weakened and the binary-check suite is closer to a valid preference proxy than this paper claims.

### 5.2 The Reasoning/Assembly Agent Distinction

The final agent in every pipeline is an assembly agent: it takes upstream outputs and arranges them into the clinical note. We found this agent type cannot be improved through prompt engineering.

Four consecutive optimization iterations. Every edit destabilized something. Fix section headers, break content merging. Preserve billing language, break duplicate detection. 18–28 point drops per attempt.

The explanation: assembly is a deterministic task assigned to a generative model. Same prompt, same input, different formatting every pass. The model treats arrangement as generation, introducing variation where none is wanted.

The fix: move all assembly quality to deterministic post-processing. Stop optimizing the assembly agent. At the time of the headline metrics we ran 43 repair rules on the assembly agent alone, more than any other agent in the pipeline. Six weeks later we did something stronger: we removed the LLM call from chart assembly entirely (§6).

**An observation for multi-agent systems:** in our pipeline, reasoning agents responded to prompt engineering while assembly agents did not. If this pattern holds more broadly, multi-agent optimization frameworks should distinguish between agents that generate new content from context and agents that arrange existing content into a format, applying prompt optimization only to the former.

We are not aware of prior work identifying this distinction in the prompt optimization literature. DSPy [Khattab et al., 2023], MIPRO, and related frameworks treat all pipeline stages as uniformly prompt-optimizable. Our evidence from one production pipeline suggests this assumption may be incorrect for assembly-stage agents, though replication across other pipelines and domains would be needed to confirm generalizability.

#### 5.2.1 Construct validity

“Assembly agent” is defined here operationally, it is the agent on which prompt optimization repeatedly regressed and which we eventually replaced, not by an a priori characterization. On a single pipeline this is unfalsifiable, and we own that.

Three registered predictions test the distinction on data we will collect: (i) **cross-profile**, four-iteration prompt optimization on the chart-assembly agents of our other five profiles should show comparable 18–28-point regressions; (ii) **replacement-intervention**, F17-style deterministic replacement on those profiles should produce comparable defect-free lifts; (iii) **counter**, if the same optimization pattern instead regresses the *reasoning* agents at ≥ 15-point magnitude, the distinction is falsified for our pipeline. Timeline: Q3 2026 cross-profile experiment.

### 5.3 Physician Preference Is Not a Single Surface

We expanded quality calibration from one rater to three board-certified physicians. Same 36 blind A/B note pairs. 84 total ratings (Table 2).

**Table 2:**
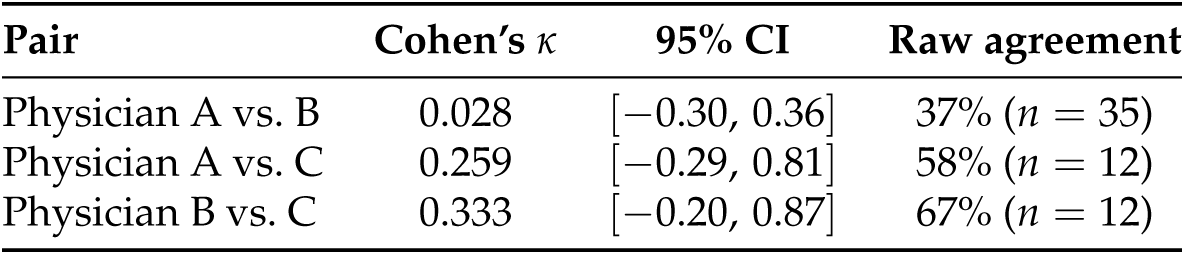
Inter-rater agreement between three physicians on 36 blind A/B note pairs. 95% CIs use the approximation defined in §5.

| Pair | Cohen’s $\kappa$ | 95% CI | Raw agreement |
| --- | --- | --- | --- |
| Physician A vs. B | 0.028 | $[-0.30, 0.36]$ | 37% ($n = 35$) |
| Physician A vs. C | 0.259 | $[-0.29, 0.81]$ | 58% ($n = 12$) |
| Physician B vs. C | 0.333 | $[-0.20, 0.87]$ | 67% ($n = 12$) |

The lower bound of each 95% CI is at or below zero, meaning that at this sample size the data are consistent with no agreement above chance for all three pairs. This is the structural finding: *n* = 35 is the largest cell, and even there the data cannot rule out *κ* = 0. We do not interpret the point estimates as ordering the three physicians’ calibration; we interpret the lower bounds as showing that all three pairs are underpowered.

At this sample size (*n* = 35 overlapping pairs for the primary rater pair, *n* = 12 for the two secondary pairs), the *κ* estimates carry wide confidence intervals. We report this as below the typical range reported in clinical documentation quality studies but directionally consistent with observations that holistic quality judgments show lower agreement than specific completeness items [Stetson et al., 2012], not as a precise population estimate. The directional finding, that two board-certified emergency physicians disagreed more often than they agreed on which note they would sign, motivated our architectural decision.

A pairwise LLM judge calibrated against one physician achieved 61% agreement with his ratings (*n* = 36). This represents single-point calibration rather than validated scalable oversight. Against the majority vote of three raters: 34%. The judge had learned one physician’s preferences, not a generalizable quality standard.

#### “Fix-verified” status is not the same as physician-preferred

Rater C’s calibration pairs compare each thumbs-down case’s original_output, the note the physician originally flagged, against a fresh generation from the current production pipeline on the same dictation. Across 12 decided ratings, rater C preferred the baseline 8 times (67%). All six distinct underlying thumbs-down cases carried status=fixed or status=closed at the time of rating. Rater C’s free-text comments cite specific, recurring issues that the failure-pattern-targeted fixes do not address. “Regression gate passed” under-constrains whether the note is actually better end-to-end. We added a physician-review queue on top of the regression gate: resolved-ticket cases are sampled into calibration pairs on a rolling basis, and a baseline-preferred vote from any qualified rater returns the case to the re-verification queue rather than letting the fixed status persist unchallenged.

This motivated the three-axis architecture and an operational definition of quality. Given that physicians disagree at near-chance levels, we do not optimize toward a single “quality” target. Instead, we define quality as a composite of three independently measurable properties that each contribute an axis to the Pareto commit rule: (1) *factual grounding*, absence of fabricated findings, verified by binary checks against the source dictation; (2) *structural/narrative completeness*, required sections, formatting, and narrative quality dimensions, enforced by deployment gates and scored per-iteration; and (3) *physician acceptance*, tracked via pairwise preference, acknowledging that acceptance criteria vary across raters.

#### Registered replication

At *n* = 120 ratings across 6 board-certified physicians on a stratified case sample, we predict *κ*_pair_ to remain below 0.30 with a 95% CI that excludes 0.5. If *κ*_pair_ *>* 0.5 at *n* = 120, the rater-fragility finding is falsified for our scale and the operational quality definition collapses to a single surface. Timeline: Q3 2026 multi-rater expansion.

### 5.4 Prescriptive Judge Instructions Regress

On this sample (*n* = 36 pairs, single-rater ground truth), the minimal instruction outperformed all three prescriptive variants (Table 3; full prompt texts in Appendix B). We hypothesize that the minimal instruction allows evaluation criteria to shift per case, matching how physicians work: a sepsis note is evaluated differently from a fall note. Prescriptive rules force fixed dimensions that may not match the evaluator’s actual decision process. A larger-scale evaluation across multiple raters would be needed to confirm this pattern.

**Table 3:** Pairwise judge variant agreement with physician preference (*n* = 36).

| Judge variant | Agreement |
| --- | --- |
| Minimal (“which would you sign?”) | 61% |
| + Signability checklist | 47% |
| + Few-shot examples | 53% |
| + Structural rules | 52% |

## 6 Architectural Follow-Through: F17 Deterministic Chart_Builder

The four observations above closed the prior version of this paper on April 17, 2026. We then operated the system through May. This section reports the architectural follow-through that the assembly-agent observation made possible.

### 6.1 Prediction, then deletion

§5.2 ended on the claim that “deterministic post-processing is an effective intervention for assembly agents.” At publication time the operational form of that claim was *rule-based repair stacked on top of an LLM-generated draft*: the assembly LLM still ran, and we wrapped 43 repair rules around its output to fix what it broke.

Six weeks later, on 2026-05-20, we deployed F17. The Chart_Builder agent on the hospitalist admission profile was migrated from a multi-kilobyte LLM prompt to a compact deterministic template (under 500 characters) plus sandboxed scripting that arranges the upstream agent outputs. The LLM call was removed entirely from chart assembly. The architectural endpoint of the assembly-agent thesis was not better wrapping of an LLM call. It was deletion of the LLM call.

### 6.2 What the deterministic Chart_Builder does (and does not do)

The new Chart_Builder consumes the structured outputs from the seven preceding LLM agents in the admission pipeline (routing, history-of-present-illness narrative, plan narrative, physical-exam narrative, summarizer, and the two assessment-and-plan agents) and renders them into the final note format using a fixed deterministic template plus a small set of sandboxed helpers. There is no generative step in assembly. The template renders sections in a fixed order with fixed transition phrasing; the helpers handle deterministic operations such as duplicate-line removal and section-header normalization. These helpers are versioned with the template, run identically on every input, and have no model parameters to drift.

Two things the new Chart_Builder explicitly does not do: it does not synthesize content from outside its upstream inputs (it cannot fabricate); and it does not paraphrase the upstream content (it renders verbatim where possible). A class of failure that previously occurred on empty or low-signal upstream input, the assembly LLM defaulting to a canonical training-data prior (e.g., 78-year-old male, pneumonia with sepsis, ceftriaxone 1g IV, bilateral infiltrates), is now structurally impossible. There is no LLM to emit a prior.

### 6.3 What it changes operationally

Three concrete operational consequences:

**(i) Latency and cost.**

We measured the F17 latency effect directly on the HM admission chat pipeline (the H+P trace in our observability platform) by comparing trace durations in the pre-F17 window (May 1–19, 2026; *n* = 100 traces) against the post-F17 window (May 20–23, 2026; *n* = 100 traces) in the same UAT environment running comparable test-bank batches (Table 4). The mean H+P latency dropped from 19.6 s pre-F17 to 10.8 s post-F17, a difference of −8.8 s (95% CI [−10.5, −7.1] s, two-sample SE-of-difference; method per §5). The standard deviation narrowed by 38% (7.4 → 4.6 s), consistent with the structural prediction: removing the LLM call eliminates a stochastic long-tail contributor, and pre-F17 traces included occasional 35–45 s outliers absent from the post-F17 distribution. The deterministic helpers themselves run server-side in single-digit milliseconds.

**Table 4:** HM admission H+P trace latency, pre- vs post-F17. Traces fetched from production observability (UAT environment); both windows ran the same batch test bank. Latencies in seconds.

| Window | $n$ | Mean | Median | SD | Min | Max |
| --- | --- | --- | --- | --- | --- | --- |
| Pre-F17 (May 1–19, 2026) | 100 | 19.6 | 17.9 | 7.4 | 6.7 | 45.0 |
| Post-F17 (May 20–23, 2026) | 100 | 10.8 | 10.1 | 4.6 | 4.5 | 31.5 |
| Difference (post – pre) | | $-8.8$ | $-7.8$ | | | |

For a separate post-F17 operational anchor on a different specialty, the currently-deployed Sayvant production pipeline averages 22.0 s end-to-end per note on the EM benchmark dataset (95% bootstrap CI [20.7, 23.4] s, *n* = 23 emergency-medicine cases); a single-call non-Sayvant scribe baseline on the same case format averages 8.8 s per note for context.

**(ii) Stochastic variance.**

Stochastic regenerations of the same note previously diverged 15–27 percentage points per-case on overall pass rate. With the assembly step deterministic, the same upstream inputs produce byte-identical chart output, removing one independent source of variance from the closed-loop commit signal.

**(iii) Safety net surface.**

The 43 repair rules that previously sat on top of the assembly LLM no longer have an LLM to repair. A subset moved upstream to attach to the LLM agents that produce Chart_Builder’s inputs. Another subset became template logic directly. A third subset, rules that existed only to clean up artifacts the assembly LLM introduced, was retired. Between the assembly-LLM removal (May 20) and the subsequent curation pass (May 21), Chart_Builder’s safety-step registry was pruned from 80 steps to 64. We then added a series of twenty new chart-quality safety nets over the following 24 hours as we surfaced edge cases in the new deterministic pipeline.

The net registry is regression-tested against the same complaint test bank that gates the rest of the loop. On a 51-case holdout drawn from production traces across the thirteen-site deployment footprint and held out from the optimizer, three sequential interventions produced a measurable lift on the defect-free rate (Table 5):

**Table 5:** Defect-free rate progression on a 51-case holdout across three sequential interventions deployed 2026-05-20. Each row is the cumulative pipeline state at that intervention; the right column is the marginal effect of adding that step alone. “F17” refers to the deterministic Chart_Builder replacement; the cleanup-step chain and dedupe step are described in §6.5.

| Stage | Intervention added | Defect-free | Rate | $\Delta$ |
| --- | --- | --- | --- | --- |
| S1 | F17 + cleanup chain in chronological order (pre-reorder baseline) | 25/51 | 49% | , |
| S2 | Reorder cleanup chain to run *after* F17 | 34/51 | 67% | +18 pp |
| S3 | Add a deduplication cleanup step | 43/51 | 84% | +17 pp |

The 49% → 84% lift over the 2026-05-20 deployment day is the composite of two interventions: (i) re-ordering the cleanup chain to run *after* F17 (§6.5 explains why) and (ii) adding a deduplication cleanup step. Holdout case-level identities are preserved across stages, but we did not record per-case pre/post status sufficient to compute paired-test discordant cell counts; we therefore report marginal rates and per-step deltas only, and avoid claiming a McNemar *p*-value we cannot reproduce. The composite +35-percentage-point improvement is the headline operational finding; the decomposition matters because it shows the F17 deployment alone (pre-S1, reported separately in our internal training-set comparison) does not account for the full lift, the post-F17 cleanup-chain ordering and the dedup step each contribute roughly equal marginal increments.

The pre/post-prune regression harness re-ran a 39-case holdout on 2026-05-21 with 39/39 cases retaining their pre-prune pass status, i.e., the curation pass that reduced 80 steps to 64 did not regress any holdout case.

### 6.4 What it does not change

F17 affects one assembly step in one profile across the thirteen-site footprint; five other profiles still wrap their assembly LLM, and the reasoning agents upstream of Chart_Builder remain LLM calls. The latency before/after is reported in §6 (Table 4); the corresponding controlled studies on physician-preference tracking and edit-distance remain future work. The scope-of-claim caveats accumulate in Limitations (§10).

### 6.5 Operational footnote: F17 reorder investigation

The cleanup steps that operate downstream of F17 (a small chain of guards targeting specific recurring defect patterns we do not enumerate here) were initially deployed in chronological order, which placed them *before* F17 in the safety-step execution sequence. Because F17 overwrites the chart with deterministic assembly from upstream agent outputs, any cleanup that ran before F17 was discarded. Reordering the cleanup chain to run *after* F17 produced the S1 → S2 lift in Table 5 (49% → 67%, +18 pp); adding a deduplication cleanup step produced S2 → S3 (+17 pp). A subsequent attempt (2026-05-21) to reorder F17 itself to run before an upstream verbatim-enforcement step destabilized two prior-passing complaint cases in the smoke gate and was rolled back. The architectural change is reported as the orderable unit; the specific position of F17 within the safety-step sequence is empirically constrained, and the ordering of downstream cleanup relative to F17 is the load-bearing detail.

## 7 Safety Architecture

The autonomous loop optimizes production clinical AI. Multiple layers prevent harm (Table 6).

**Table 6:** Safety layers in the autonomous optimization pipeline.

| Layer | Mechanism | Trigger |
| --- | --- | --- |
| Static lint | Syntax + schema + anti-pattern regex | Each iteration |
| Cross-iteration memory | Edit signatures rejected/reverted 3+ times surfaced to the agent | Each iteration (pre-agent) |
| Runtime smoke | 1-case end-to-end pipeline check; auto-revert on fail | Each iteration (post-lint) |
| Per-iteration | File allowlists, infrastructure checks | Each iteration |
| Binary gate | Gate-class fabrication checks: zero tolerance | Each iteration |
| Data isolation | File-level holdout/training split with path guards | Every data access |
| Holdout | Unseen test cases; >10% divergence flagged | After commits |
| Regression | All resolved complaints re-verified | After each commit |
| Pre/post-prune harness | Holdout re-run after safety-step registry edits | After prune/add waves |

The regression gate is the hardest constraint, implementing what Shah et al. [2023] call “recur-ring local validation”, the principle that one-time external validation is insufficient for deployed clinical AI, and that site-specific reliability tests must run before every deployment. A physician’s previously resolved complaint cannot be broken by score optimization. Physician trust compounds, regressions destroy it.

### Regulatory and ethical considerations

The system optimizes the instructions given to generation agents, not the clinical content of individual notes. No patient data enters the optimization loop: the nightly cycle operates on de-identified test cases derived from production traces. The system’s output, modified agent prompts and repair rules, affects future note generation but does not alter any existing medical record.

Whether autonomous prompt optimization for clinical documentation constitutes a modification to an AI/ML-based software as a medical device (SaMD) under the FDA’s predetermined change control plan framework is an open question. Our system modifies the behavior of a clinical documentation tool without human review of each individual change, which could fall within scope of the FDA’s proposed regulatory framework for ML-based SaMD. We the optimization targets documentation fidelity against the source dictation, it does not generate clinical decisions or recommendations. The distinction between optimizing documentation fidelity and optimizing clinical decision support may be relevant to regulatory classification but has not been formally adjudicated.

### 7.1 Operational Lessons

Five production outages in the December 2025–January 2026 window exposed a gap between quality intelligence and operational resilience. All five outages were detected manually, not by the scoring harness. All shared a root cause: deterministic repair rules (Layer 1) corrupting JSON-outputting agents, a punctuation cleanup function destroying JSON structure, a variable reference returning empty on summarizer profiles, short substring matching inside longer words, and an overly restrictive JSON schema forcing empty output.

These were not quality failures that the binary check system could catch; they were structural failures that corrupted the output pipeline itself. In response, we deployed pre-deploy circuit breakers (static analysis targeting the five known outage patterns), continuous production monitoring (4-hour trace sampling with structural validation), deploy snapshots with auto-rollback, and automated promotion gates replacing manual weekly review.

### 7.2 Observability

Four observability mechanisms define what is auditable about each run.

#### Prompt fingerprinting

Every iteration records a cryptographic hash (truncated for storage) and character length of the editable prompt surface (the agent prompt modules, the deterministic-repair profile JSON, and the output validator). Fingerprints are written to a per-iteration log alongside the commit/revert outcome. Diffing consecutive hashes across iterations surfaces silent prompt bloat, drift that does not produce a visible score change but accumulates over weeks. The combined hash also appears on pairwise history entries, giving each committed pairwise vote a precise reference to the prompt state that produced it.

#### File-level data isolation

Holdout and training data access is partitioned at the module level. A single dedicated module is the only one permitted to read from the holdout directory and refuses any path outside the holdout roots. A mirror module on the training side refuses any path containing the holdout marker. This physical separation enforces at the file level what an opt-in flag would only request procedurally, eliminating a class of accidental contamination.

#### Separate holdout log

Holdout evaluation results are appended to a dedicated file that the holdout-access module is the only writer of. This log functions as a reproducibility receipt: the holdout file is both isolated at read-time and isolated at write-time.

#### Skipped-iteration logging

Iterations rejected by self-review or the runtime smoke gate previously left no entry in the iteration log, only committed and reverted iterations were persisted. The current implementation records these as action: skip or action: revert with full fingerprint attached, producing a complete audit trail of every edit the agent proposed.

#### Per-type domain specifications

Each note type (admission, progress, discharge) has a single-file canonical specification that names the unit of evaluation, the fixed base model, the success metric, the split policy, the harness boundary (which files the optimizer may edit), and the per-run budget. The specifications are the authoritative per-type scope.

### 7.3 Production Operational Metrics

A closed-loop quality system cannot be fully evaluated on intrinsic loop metrics alone; deployment-outcome metrics live downstream of the optimizer in production usage. Five categories are operationally relevant: physician time-to-sign, median edit distance between generated and signed notes, per-agent regeneration rate, post-deployment complaint re-submission rate, and end-to-end latency distribution per note type. Of these, latency is instrumented (the Sayvant production pipeline emits per-agent duration via Langfuse traces); the others are tracked in the deployed-customer billing and EHR-integration surface and are not in a form we can report in this paper. We flag this as a structural gap: the architecture argued for in §5.2–§6 (deterministic assembly removes a stochastic latency contributor and a generative variance source) should be evaluated end-to-end on the metrics above. That evaluation is in progress and will be reported separately.

## 8 Results

The 39% keep rate is by design. The safety system rejects most candidate changes, preferring zero improvement over any risk of regression (Table 7). Nightly runs cost approximately $3.50 each, with the Claude optimizer running on a flat-rate subscription and binary check scoring on a separate per-token basis. The 39% keep rate is an aggregate across the pre-Pareto single-signal regime; per-iteration prompt fingerprinting began 2026-04-16 and enables longitudinal trend analysis in subsequent reports.

**Table 7:** System operational metrics (36 days, March–April 2026, pre-Pareto single-signal regime). Numbers in this table predate the F17 architectural follow-through (§6).

| Metric | Value |
| --- | --- |
| Nightly runs | 135 |
| Total optimization iterations | 1,089 |
| Committed improvements | 290 (39% keep rate)<sup>†</sup> |
| Reverted candidates | 453 |
| Failure patterns cataloged | 226 |
| Patterns resolved | 161 (71%) |
| Deterministic repair rules | 166 across 6 profiles |
| Binary evaluation checks | 282 (59 macro, 223 micro) |
| Agents maintained | 66 across 4 note types |
| Prompt codebase | 40,114 lines |
| Physician complaints tracked | 42 unique |
| Regressions on tracked complaint cases | 0 |
| SG pass rate improvement | +11.7 pts ($n = 25$) |
| SG fabrication elimination | 6 → 0 failures ($n = 25$) |
| *Cost* |  |
| Nightly optimization run | ~\$3.50 (~3 hours runtime) |
| Binary check scoring per note | ~\$0.10 |
| Full pipeline per admission note | ~10 minutes (6–19 agents) |

<sup>†</sup>The 39% keep rate is the aggregate across the pre-Pareto *single-signal* commit rule (“binary target must improve; failures cannot rise by more than 2”). The three-axis Pareto rule described in §3 replaced it on 2026-04-16; Pareto-era keep rate is reported as ongoing and is expected to be lower because the rule rejects more candidates.

### 8.1 Ablation: Contribution of the Deterministic Repair Layer

To quantify the contribution of each system layer, we evaluated 25 production test cases under three conditions using the full 282-check binary evaluation matrix (Table 8).

**Table 8:** Ablation results: 25 test cases evaluated against 282 binary checks under three conditions. “Checks defect-free” is the proportion of the 282 checks that pass on *all* 25 cases. “Per-case pass” is the mean of the 25 case-level pass rates (each case’s count of passing checks ÷ 282). CI methods per §5.

| Condition | Checks defect-free | Per-case pass | Failing | Fab. |
| --- | --- | --- | --- | --- |
| C: Bare pipeline (no repair) | 68.8% [63.2, 73.9] | 94.3% [92.7, 95.9] | 88 / 282 | 6 |
| B: Server-side SG only | **80.5%** [75.5, 84.7] | **95.9%** [94.7, 97.0] | 55 / 282 | **0** |
| A: SG + client-side repair | 58.9% [53.0, 64.5] | 91.6% [88.6, 94.6] | 116 / 282 | 0 |

#### Server-side deterministic repair (SG) contributes +11.7 percentage points on the checks-defect-free metric

(Condition C → B. Wilson CIs above), driven primarily by the elimination of all 6 fabrication-class failures (6 → 0). The fabrication failures in Condition C include fabricated pertinent negatives, fabricated medication orders, and fabricated code status language, exactly the categories the repair rules target.

The per-case pass rate tells a more conservative story: paired-bootstrap on the 25 per-case rates puts the B − C improvement at +1.6 percentage points with 95% CI [−0.2, +3.5]. The CI crosses zero. At *n* = 25 cases, the per-case mean cannot reject the null of no improvement, even though the checks-defect-free metric shows a clear gap. This is the honest description: SG repair eliminates specific high-severity failure modes (fabrication 6 → 0; checks-defect-free +11.7 pp), and at this sample size we cannot also claim that aggregate per-case quality improves. The destructive effect of client-side repair is reliable on both metrics (per-case B − A delta = −4.4 pp, 95% CI [−7.4, −1.5]).

A supplementary omission analysis on a 5-case subsample categorized each of the 282 checks as fabrication (17), omission (180), or other (85). Across all three conditions, fabrication failures were near-zero (≤1). Omission failures, by contrast, increased slightly from bare pipeline (35) to SG-repaired (40) to client-repaired (44), suggesting that repair rules that suppress fabricated content may inadvertently remove legitimate content in the process. The additional omissions introduced by repair rules are predominantly in the major-severity category, suggesting the fabrication-omission tradeoff involves clinically significant content. This warrants further investigation at larger sample sizes with explicit per-case severity annotation.

#### Client-side repair is destructive

The client-side autoRepairOutput() layer, originally designed before the binary check suite existed, reduces pass rate by 21.6 points (Condition B → A). Investigation shows it aggressively strips content that binary checks expect to be present, over-correcting fabrication at the cost of completeness. This is a degenerate feedback loop: an autonomous repair system that reinforced the wrong signal because its evaluation criteria predated the binary check suite. This layer has been disabled.

This result supports the architectural claim that deterministic post-processing is an effective intervention for fabrication-class errors in our pipeline. It also demonstrates the value of ablation, without this comparison, the destructive client-side layer would have remained undetected. The May 2026 follow-through (§6) extends the argument: replacing the assembly LLM call with a deterministic renderer is a stronger version of the same intervention.

The system catalogs failure patterns faster than it resolves them (226 cataloged, 161 resolved) because physician complaints and production traces continuously surface new patterns. The 16 complaints verified fixed autonomously (of 42 total) reflects a conservative pipeline: many complaints map to known limitations rather than fixable prompt or repair rule issues.

The closest published comparator is Chen et al. [2026], who deployed an agentic LLM discharge summarizer with 11 hospitalists and found a 2% hallucination rate but 25% omission rate and 20% inaccuracy rate, demonstrating that even carefully engineered clinical AI systems require post-generation quality assurance. Production clinical AI systems (Nuance DAX Copilot, Abridge, DeepScribe) have not published closed-loop autonomous quality assurance architectures.

## 9 Related Work

### LLM-as-judge evaluation

Recent work shows that LLMs can evaluate generated text effectively [Zheng et al., 2024, Kim et al., 2024]. The closest lineage for the pairwise judge described here is the family of preference-learning approaches developed for RLHF, Christiano et al. [2017], Ouyang et al. [2022], and the constitutional / RLAIF variant that scales preference signal via model judgments [Bai et al., 2022]. Our judge differs in two ways: it is a calibration target rather than a training signal, and it scores against a single physician’s preferences rather than an aggregated population reward model. Our observation that minimal pairwise instructions (61% agreement) outperformed three prescriptive variants (47–53%) on *n* = 36 pairs is, to our knowledge, a novel finding in clinical documentation evaluation. We hypothesize this reflects the case-specific nature of clinical quality judgments, though larger-scale replication is needed.

### Self-refinement and iterative improvement

The nightly loop is structurally adjacent to single-session self-refinement systems, Madaan et al. [2023] SELF-REFINE and Shinn et al. [2023] Reflex-ion, which use LLM critics to revise outputs within a single inference chain. Our loop differs in operating across iterations on the *prompts and rules* that produce outputs, rather than on outputs themselves, and in gating each iteration through a multi-axis Pareto rule with absolute-floor reverts. The cross-iteration rejection memory (§3.5) is adapted from a similar replay-protection pattern in HuggingFace’s ml-intern doom_loop.py.

### Inter-rater agreement

Low inter-rater reliability is well-documented in clinical quality assessment. ICD-10 coding studies report mean *κ* = 0.67 overall, with 4-digit specificity dropping to *κ* = 0.2–0.4 [Cheng et al., 2024]. Chart-review reliability studies more broadly report substantial variation depending on rater training and instrument structure [Lorenzetti et al., 2018]. Stetson et al. [2012] developed the PDQI-9 instrument and reported good inter-rater reliability (ICC = 0.83), though subsequent applications in less controlled settings have shown wider variation. Our *κ* = 0.028 is below the typical range reported in this literature (*κ* = 0.3–0.7 for clinical note quality audits), making it an outlier rather than a typical finding, though it is directionally consistent with observations that overall quality judgments show lower agreement than specific completeness items.

### Autonomous prompt optimization

DSPy [Khattab et al., 2023], TextGrad [Yuksekgonul et al., 2024], and OPRO [Yang et al., 2024] optimize prompts against fixed metrics. Our contribution: the metric matters more than the optimizer. We further observe that assembly-stage agents in our pipeline did not respond to prompt optimization (§5.2), and that the strongest version of the architectural response is to replace the assembly LLM call entirely (§6), a distinction we have not found discussed in this literature. Critically, none of these systems operates in production on safety-critical outputs across multiple deployment sites with real physician feedback; the constraints that drove our acceptance rule (Pareto-with-absolute-floors) and our cross-iteration rejection memory are downstream of that operational setting and do not arise from a pure-optimization framing.

### Production clinical AI deployment

Operational writeups of deployed clinical AI emphasize that the bulk of the work is post-modeling: Sendak et al. [2020] describe the deployment of a sepsis-prediction model and frame the gap between offline accuracy and operational outcomes as the dominant deployment challenge. Sculley et al. [2015] characterize the long-tail engineering debt of ML systems in production: glue code, monitoring gaps, undeclared consumers, and feedback loops. The closed-loop architecture in this paper attempts to address that debt explicitly for the clinical documentation case, with the safety architecture (§7) and observability mechanisms (§7.2) targeted at the system-level failure modes those authors describe.

### Post-hoc output repair

Rule-based post-processing of clinical NLP output has a long lineage: Friedman et al. [2004] (MedLEE) and Savova et al. [2010] (cTAKES) both apply layered rule systems on top of statistical extraction pipelines to enforce semantic constraints and reconcile output against source text. What we add is not the use of rules per se but the density (initially 166 rules across 6 deployed profiles, then restructured around a deterministic renderer for the chart-assembly step), the integration into an autonomous nightly optimization loop, and the source-dictation cross-referencing at the LLM-pipeline-stage granularity rather than at the corpus-extraction layer. Constrained decoding is a generation-time alternative that cannot use source-document comparison; the post-hoc approach here is complementary, not competing.

## 10 Limitations

The inter-rater study is underpowered (*n* = 3 raters, *n* = 36 pairs, one rater contributed only 9 ratings). The *κ* estimates carry wide confidence intervals at these sample sizes. The *κ* = 0.028 finding motivates our architecture but should not be generalized without larger-scale replication across more raters and sites.

The system operates on one organization’s multi-agent pipeline (Sayvant Health) deployed across thirteen US hospital sites; generalization to other vendors’ pipelines, to international clinical settings, to federal/VA sites, and to pediatric-only institutions is untested. The reasoning/assembly agent distinction (§5.2) is observed from four optimization attempts on one assembly agent within this pipeline, replication across other multi-agent pipelines (DSPy-compiled programs, other commercial clinical-documentation vendors) is needed before this can be considered a general principle. The F17 architectural follow-through (§6) is one assembly agent on one note type; five other profiles still run an assembly LLM with repair-rule wrapping at the time of writing.

The fabrication elimination (6 → 0) comes from 25 test cases drawn from the multi-site case pool; the ablation is not powered for per-site stratified generalization. The system evaluates final note quality but does not separately measure intermediate-stage accuracy. Binary checks are authored by the same team operating the system, creating potential confirmation bias in what is measured. The 282-check suite has not been independently validated by external reviewers (see audit pre-registration below).

The system’s human feedback signal (42 physician complaints over 36 days) is thin compared to recommendations of 30–1,000 daily human-evaluated examples for production AI systems. The system does not measure patient outcomes, physician time-to-sign, or note edit rates. Quality is defined operationally (§5.3) in terms of fabrication absence, structural completeness, and physician preference, not clinical outcomes. Whether improved documentation fidelity translates to improved care remains an open question.

The system raises a deskilling concern that we do not currently measure. As AI-generated notes become the default starting point, physician skill at composing a complete note from scratch can atrophy in ways that are invisible when the AI substrate is working and severe when it fails. The system commits prompt changes to production clinical documentation tools autonomously, with human oversight limited to veto windows and Slack notifications. Whether technical safety gates (binary checks, regression protection) constitute sufficient institutional governance for autonomous modification of clinical AI tools, as distinct from per-case “human in the loop” oversight, is an open question that extends beyond regulatory classification into institutional AI governance [Haque et al., 2020, Stanford HAI, 2022].

Institutional disclosure norms for autonomous optimization of clinical documentation AI, as distinct from disclosure of AI use in documentation itself, are unsettled, and the system described here would benefit from such norms being articulated prospectively rather than retroactively.

### Independent check-validity audit (planned)

The 282 binary checks reported in §3 and the ablation in §8 were authored by the same engineering team that operates the closed-loop system. This is the most consequential confirmation-bias risk in the paper: the system optimizes against checks the same team designed, then reports passing rates on those checks as evidence of quality. To mitigate, we are committing prospectively to a 30-case external-rater check-validity audit before this paper’s submission to peer review. The protocol: a stratified random sample of 30 binary checks (drawn proportionally across the six rule categories in Table 1), independently reviewed by two non-team clinicians who have not contributed to the system’s development; each check is rated for face validity (does this check measure a clinically meaningful property?) and for construct relevance (is the failure mode this check detects actually a documentation defect rather than a stylistic preference?). The audit instrument, the resulting agreement statistics, and any checks failing either criterion will be reported in an addendum prior to peer-review submission. We pre-register the threshold for action: any check failing either criterion from ≥ 1 rater will be removed from the optimization target and the ablation will be re-computed.

### Multiple-comparisons disclosure

This paper reports several inferential statements (Spearman *ρ* in §5.1; three Cohen’s *κ* values in §5.3; per-case bootstrap delta in §8) without family-wise multiple-comparison adjustment. We report per-test 95% CIs and treat individual results as descriptive within the small-*N* regime; we do not claim joint significance. Readers comparing these results to a higher evidentiary bar (e.g., a family-wise Bonferroni-adjusted *α* = 0.05/*k*) should several of the reported borderline findings would no longer cross significance under that adjustment.

### Downstream stakeholders not directly evaluated

The system’s outputs are downstream-consumed by clinical-documentation-improvement (CDI) specialists, medical coders, and quality-review teams whose perspective on note quality may diverge from physician-acceptance preference. We have not surveyed those stakeholders. CDI specialists in particular evaluate notes against billing-compliance criteria (correct MCC capture, diagnostic specificity, present-on-admission flags) that overlap with but are not identical to the physician-signability axis used by the pairwise judge. A complete operational evaluation would include CDI/coder-rater inter-group reliability against the same case set; this is future work.

### Human-AI teaming and physician deskilling

The deskilling concern in §5.3 sits within a broader literature on physician interaction with deployed clinical AI [Topol, 2019, Patel et al., 2019]. Topol [2019] argues that AI-augmented documentation could either return physician attention to the patient or further atrophy clinical-narrative skill, depending on whether the AI substrate is treated as a draft to be edited or a finished product to be signed. Our system is currently treated as the latter at the deployed site, the physician edits in Epic after Sayvant generates the note, and the autoresearch loop optimizes against the pre-edit note. Per-physician edit-rate measurement (deferred to §7.3) is the load-bearing missing observation here: a chronically-falling edit rate over time would be consistent with both improvement (AI gets better) and atrophy (physician stops checking).

## 11 Conclusion

Quality maintenance for deployed clinical AI is an operational problem, not a modeling problem. The bottleneck is not generating better notes, it is finding problems, diagnosing their root cause, fixing them without breaking other cases, and proving the fix works. At scale, this requires automation.

We present a closed-loop system where physician complaints drive test case creation, autonomous overnight optimization generates and evaluates fixes, multiple safety layers gate deployment, and physicians are notified when their complaints are resolved. Four observations from our system may generalize.

First, the same binary-check instrument produces opposite outcomes depending on the question asked: “increase this score” produces structurally-correct notes that physicians reject; “did this specific fabrication stop?” produces rater-invariant deployment decisions. The instrument is the same; the framing determines the result.

Second, in our pipeline, assembly-stage agents did not respond to prompt optimization the way reasoning agents did. We hedge this as a hypothesis pending the three falsifiable predictions registered in §5.2.1, but the operational evidence is strong enough that six weeks after the prediction we replaced the assembly LLM call entirely.

Third, physician preference is rater-fragile at samples sizes typical of clinical-AI calibration studies. Cohen’s *κ* = 0.028 with 95% CI [−0.30, 0.36] on *n* = 35 overlapping pairs does not establish disagreement; it establishes that single-rater calibration is structurally insufficient. The three-axis Pareto rule with absolute-severity floors is our operational response to this fragility.

Fourth, and this is the architectural punchline, when prompt optimization fails on a pipeline stage, the strongest version of the intervention is not better post-processing wrapped around the LLM call. It is deleting the LLM call. F17 removed the assembly LLM from a six-to-nineteen-agent pipeline and replaced it with a compact deterministic renderer. Deterministic post-processing was a way station; the endpoint was structural.

These observations come from one system (Sayvant Health) deployed across thirteen US hospital sites over a 60-day window, with the F17 follow-through reported through May 2026. The thirteen-site footprint spans academic-to-community variation in patient population, payer mix, and staffing model; whether the observations replicate across *other vendors’* clinical AI systems is the empirical question we hope this work motivates others to investigate. The system’s architectural contributions, including the closed-loop architecture and the Pareto-with-absolute-floors acceptance rule, are enumerated in §11; this conclusion is intended to characterize the *findings*, not to re-enumerate them.

## Reproducibility and what is withheld

The numerical results in this paper (Tables 3, 2, 8, 7) are reproducible from a bundle of anonymized ablation per-case counts, bootstrap-CI data, and analysis scripts, released under CC BY 4.0 at https://github.com/sayvant/SQS-Auditor-paper-data/releases/tag/paper-arxiv-v1.0-data. The bundle includes reproduce_ablation_ci.py (Python 3.11, seed 42, 10,000 bootstrap re-samples) which reproduces the Table 8 CIs exactly. A Zenodo DOI for the same bundle will be registered prior to peer-review submission. The four pairwise-judge prompt variants are described structurally in Appendix B (per-variant structure plus reported agreement against single-rater ground truth); their verbatim text is withheld under category (iii) below.

Five categories of material are withheld from the public release for operational-security, site-confidentiality, and competitive-protection reasons: (i) the content of the deterministic repair-rule library and the full enumeration of the binary check suite; (ii) per-axis numerical thresholds in the Pareto commit rule and the severity-classification decision tree referenced in §3.3.1; (iii) the internal prompt text of the reasoning and assembly agents in the production pipeline, including the verbatim text of the four pairwise judge variants whose structure is described in Appendix B; (iv) the specific identities of the thirteen US hospital sites in the deployment footprint, including per-site case attribution for any test-bank or ablation sample; (v) the implementation surface of the closed-loop system (source-tree file paths, function names, internal versioning identifiers for individual safety steps beyond the named F17 milestone). None of these is load-bearing for the observations reported: the patterns concern the *shape* of the commit rule, the structure of the evaluation surface, and the existence of the architectural decisions, not the specific rule content, exact wording, site identities, or implementation surface. Researchers interested in replicating the harness at a different site can do so with their own rule library; researchers interested in replicating the patterns can do so from the architectural description and the released aggregate-result files alone.

## The reproducibility-vs-IP trade-off, made explicit

The withholding scope above is larger than the in-paper architectural description, and we want to name that asymmetry rather than hide it. We aim to describe the architectural patterns at a granularity sufficient for them to be re-implemented at a different vendor’s pipeline, the three-layer structure, the Pareto-with-absolute-floors acceptance rule, the cross-iteration rejection memory, the structural shape of the pairwise judge variants, and the deletion-of-LLM-call follow-through, but the released bundle alone is not sufficient to rebuild *this* deployed system. Replication of the *patterns* (do they hold on another vendor’s pipeline?) is what we hope this paper motivates; replication of *Sayvant’s specific implementation* is not the goal of the release. Norms for resolving this trade-off in clinical-AI deployment-science publishing are unsettled; we treat this paper as one explicit data point on where the line currently sits, and we welcome feedback from venue editors and the broader community on whether to draw it differently.

## Author Contributions

Using the CRediT (Contributor Roles Taxonomy) framework: **A.N.**, conceptualization, methodology, software (closed-loop architecture, scoring intelligence, gate hierarchy, autoresearch loop), formal analysis, investigation, data curation, writing (original draft, review and editing), visualization, supervision, project administration. **J.W.**, software (Sayvant platform. Sayvant Guard execution sandbox, profile-deployment infrastructure), resources, writing (review and editing). **M.H.**, validation (clinical guidance via the Heslin mandates registry that overrides other clinical sources for HM-specific guidance), writing (review and editing). All authors read and approved the final manuscript. A.N. accepts responsibility for the integrity of the data and the accuracy of the data analysis. CRediT role assignments have been confirmed directly with each co-author.

## Disclosures and Conflicts of Interest

### Affiliations and relationships

A.N. is a graduate student at Stanford School of Medicine (MCiM program) and is a co-founder of Sayvant Health, holding both equity and a clinical-AI leadership role at the company. M.H. is a hospitalist at Stanford School of Medicine in the same MCiM program, and serves as a paid clinical consultant to Sayvant Health; he is not a Sayvant employee. J.W. is an employee of Sayvant Health.

### Independence of analysis

The closed-loop quality system described here operates on Sayvant’s production pipeline; we describe it from the position of operating it, not from the position of an independent audit. The 282 binary checks and the deterministic repair rules in the ablation were authored by the engineering team operating the system; the planned 30-case external-rater check-validity audit (§10) is the mitigation for this confirmation-bias risk. A.N. and M.H. acknowledge that, as Stanford-affiliated researchers, they remain commercially interested in Sayvant’s success, and readers should weight the operational claims appropriately.

### Reproducibility

The numerical results in Sections 8–6 are reproducible from the anonymized rating files released alongside this paper at the public reproducibility bundle (§11). The architecture and operational findings are observed across a thirteen-site deployment footprint over a 60-day window and should be replicated by other vendors’ clinical-AI systems before being treated as universal.

### Human-subjects research status

This work analyzes the behavior of a deployed clinical AI documentation system, specifically, model outputs, structural checks on those outputs, and physician-feedback signals (thumbs-down flags) on signed notes. The analyses do not involve patient-identifiable data, do not access or use protected health information at the deployment sites, and operate on aggregate logs of system behavior rather than on individual patient records. The work was conducted at Sayvant Health and at customer deployment sites as operational engineering measurement. This study was reviewed by the Heartland Institutional Review Board (HIRB), a federally-registered IRB (FWA00023407, IRB00007694), and determined to be *exempt* from IRB review under 45 CFR 46.104(d)(8) (Secondary Research with Broad Consent) as HIRB Project No. 052726-1372, approved 27 May 2026. Per Heartland’s requirement: *this project has been deemed exempt by the Heartland Institutional Review Board.* The approval letter is included with the reproducibility bundle described in §11.

## Data Availability

A reproducibility bundle of anonymized ablation per-case counts, bootstrap-CI data, and analysis scripts is released under CC BY 4.0 at https://github.com/sayvant/SQS-Auditor-paper-data/releases/tag/paper-arxiv-v1.0-data. The deterministic repair-rule library content, full enumeration of the 282 binary checks, per-axis numerical thresholds, internal agent prompt text, site identities, and source-code implementation surface are withheld for operational-security and competitive-protection reasons as documented in the manuscript's "Reproducibility and what is withheld" section. None of the withheld material is load-bearing for the observations reported.

## Acknowledgments

We thank the board-certified physicians who contributed blind A/B ratings to the multi-physician calibration dataset, spanning two specialties across academic and community practice settings. We thank the deploying clinicians at the thirteen sites whose physician-feedback signals drove the test-case bank. The AI Council review process, a structured internal critique panel modeled on adversarial peer review, contributed material revisions to earlier drafts.

## A Gate Architecture Summary

This appendix is intended as the audit surface a deploying site would need to replicate the closed-loop architecture: every gate that sits between an autonomous edit and a production change is listed, in one table, with its role and the signal it produces. A reader implementing the architecture at another site should be able to use Table 9 as a checklist of components that must be present (or deliberately omitted with a reasoned waiver) before any autonomous commit is enabled.

**Table 9:** Consolidated gate hierarchy. “Floor” = absolute-floor revert that fires regardless of other axes. “Axis” = one input to the Pareto commit rule. “Operational” = post-commit production safety.

| Gate | Role | What it checks | Produces |
| --- | --- | --- | --- |
| TS syntax lint | Runtime | tsc -noEmit on diff | accept/reject |
| JSON validation | Runtime | profile JSON parses | accept/reject |
| Anti-pattern lint | Runtime | 5 known-bad regex patterns | accept/reject |
| Self-review | Runtime | LLM critic vs. top-3 failing checks | accept/reject |
| Smoke gate (1-case) | Runtime | end-to-end pipeline on 1 case | accept/revert |
| Binary target axis | Axis | pass rate of pattern-mapped checks | numeric delta |
| Narrative axis | Axis | 6 narrative quality dimensions (mean) | numeric delta |
| Pairwise axis | Axis | “which would you sign?” judge | vote counts |
| Pareto rule | Decision | dominance / non-dominance across axes | commit/revert |
| Gate-class floor | Floor | fabrication check $\geq 1 \rightarrow 0$ passing | force revert |
| Major-severity floor | Floor | new major-severity check fails | force revert |
| Severity-weighted pairwise floor | Floor | baseline preferred + major-severity case | force<br>pairwiseOutcome = loss |
| TD regression gate | Floor | tracked complaint breaks | force revert |
| Holdout gate | Deployment | pass rate divergence > 10% vs. training | flag overfit |
| Pre/post-prune harness | Deployment | holdout re-run after safety-step prune | flag regressions |
| Deploy snapshots | Operational | auto-rollback on production deviation | rollback |
| Production monitor | Operational | 4-hour trace structural sampling | alert |

**Table 10:**
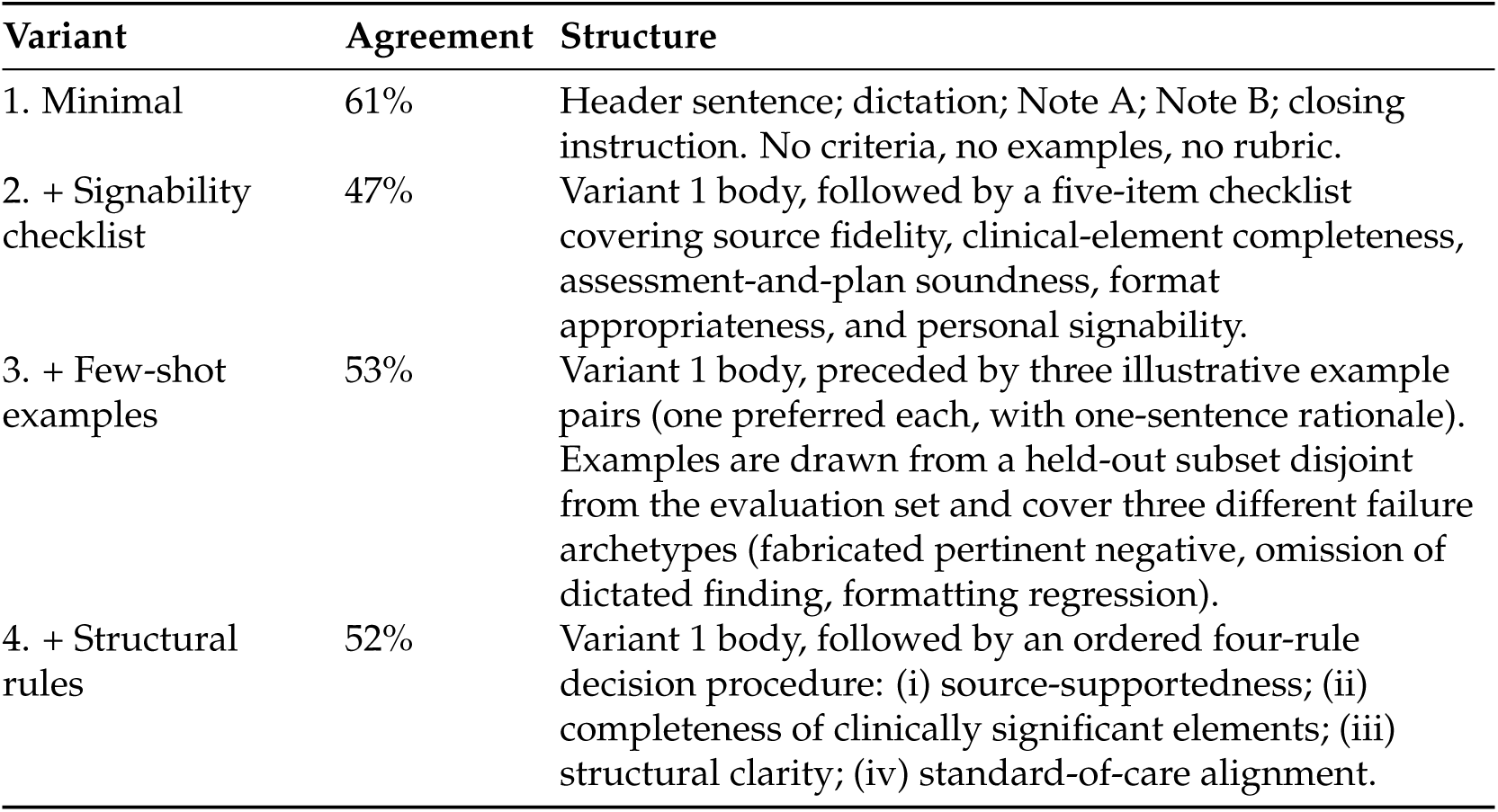
Structural description of the four pairwise judge variants. “Closing instruction” is the final imperative asking for an A/B label; this is identical across variants. Differences are in the guidance that precedes the closing instruction.

**Table 11:** TRIPOD-LLM item coverage in this paper. “Reported” = item is addressed in the body. “Deferred” = instrumentation in progress; tracked in §7.3.

| TRIPOD-LLM item | Status | Where |
| --- | --- | --- |
| Study objective and primary contribution | Reported | Abstract; §1 |
| Data sources and provenance | Reported (qualitative) | §4 (harvest step) |
| Model architecture (the pipeline being optimized) | Reported | §3, Fig. 2 |
| Training/optimization procedure | Reported | §4 |
| Evaluation methodology and metrics | Reported | §3 (Layers 2/3); §8 |
| Inter-rater agreement (where applicable) | Reported | §5.3; Table 2 |
| Operational safety architecture | Reported | §7; Appendix A |
| Bias and fairness assessment | Partial | §5.3 (rater fragmentation) |
| Per-note latency distribution | Reported | §6, Table 4 |
| Median edit distance generated → signed | Deferred | §7.3 |
| Per-agent regeneration rate | Deferred | §7.3 |
| Post-deployment complaint re-submission rate | Deferred | §7.3 |
| Limitations and harm mitigations | Reported | §7; Limitations |
| Reproducibility statement | Reported | §11 |
| Disclosure of AI assistance in writing | N/A (no AI in writing this paper) | , |

Table 9 consolidates every gate described in the body of the paper into a single reference. Three columns are noted explicitly: whether the gate is a *deployment gate* (runs between edit and production) or a *runtime gate* (runs inside the nightly loop before or during evaluation), what it measures, and what it produces (a boolean accept/reject, a signal fed to another rule, or a diagnostic log).

## B Pairwise judge prompt variants

All four variants were evaluated against the same *n* = 36 single-rater ground truth with the same model, temperature, and input format. The inputs in each case are the physician’s original dictation followed by Note A and Note B, with A/B labels randomized per pair. Each variant returns the label of the preferred note. We describe the structure of each variant; the exact prompt text is withheld for the reasons documented in §11.

All four variants received identical input in terms of dictation text, note text, and pair ordering. Differences in agreement cannot be attributed to input conditioning. Showing the dictation alongside both notes was not a variant condition, it was constant across all four. The largest single-intervention improvement we observed during judge design was the inclusion of the dictation (relative to pairwise-only comparisons without source text); that effect sits upstream of the variants reported here.

*Why the exact prompts are withheld*.

The minimal variant in particular is short, self-contained, and directly transferable; releasing it verbatim would substantially reduce the work required for a competing clinical-AI vendor to reproduce our calibration scheme without the rest of the system context. The structural descriptions above (and the agreement percentages) are sufficient for a researcher to reproduce the *finding*, that minimal pairwise instructions outperformed three prescriptive variants on this dataset, without our exact wording.

## C TRIPOD-LLM Reporting Items

The TRIPOD-LLM reporting framework [Gallifant et al., 2025] extends TRIPOD+AI to LLM-based clinical prediction and generation systems. The table below maps each TRIPOD-LLM item to the section in this paper where the item is addressed, and flags items where the corresponding instrumentation is not yet in place. Items flagged *(deferred)* are tracked in §7.3 as production-side metrics that live downstream of the optimization loop.

*Note on deferred items*.

The remaining deferred items all measure properties of the deployed pipeline rather than properties of the nightly optimizer. The latency before/after on HM admission is reported in §6 (Table 4); per-physician edit-rate, regeneration rate, and complaint re-submission tracking remain pending instrumentation.

## D Glossary and notation

This appendix collects the architectural terms used throughout the paper that may benefit from a short definition. Specific implementation-level identifiers (file paths, function names, internal versioning) are deliberately not enumerated here; see §11 for the scope of what is withheld.

### F17

The single named architectural milestone in this paper: the deterministic chart-assembly step that replaced the chart-assembly LLM call on the hospitalist admission profile on 2026-05-20. The replacement consists of a compact deterministic renderer (under 500 characters) plus a small set of sandboxed helpers that consume the structured outputs of the preceding LLM agents and produce the final note. The architectural follow-through reported in §6. We retain “F17” as a stable reference label so the paper has a single, citable name for the milestone; other internal step identifiers are referenced descriptively (“cleanup chain,” “deduplication step,” etc.) rather than by name.

### Chart_Builder

The chart-assembly agent of the hospitalist admission pipeline. Pre-F17, an LLM agent with a multi-kilobyte prompt. Post-F17, a deterministic renderer with no LLM call.

### Sayvant Guard (SG)

The deterministic post-processing layer described as “Layer 1” in §3. Executes in a sandboxed scripting environment after each LLM agent.

### Profile

A deployed configuration of one note type (e.g., admission, progress, discharge) consisting of an ordered chain of LLM agents plus their associated SG steps.

### TD-NNN

Thumbs-down identifier. Physician-flagged production case with sequential numeric ID. TD-077 is the worked example in §2; we use it as a stable reference label and do not enumerate the full TD case list. The 42 tracked complaints in this paper are all TD-identified cases.

### Autoresearch loop

The nightly closed-loop optimization process described in §4. Implementation is internal.

### SG step ordering

Within a profile, SG steps execute in a fixed sequence after the LLM agent’s output is produced. The ordering of steps relative to F17 is empirically constrained (§6.5).

